# Deep sequencing of early T stage colorectal cancers reveals disruption of homologous recombination repair in microsatellite stable tumours with high mutational burdens

**DOI:** 10.1101/2022.03.02.22271810

**Authors:** Jun Li, Pascal Steffen, Benita C. Y. Tse, Mahsa S. Ahadi, Anthony J Gill, Alexander F. Engel, Mark P. Molloy

## Abstract

Early T stage colorectal cancers (CRC) that invade lymph nodes (Stage IIIA) are greatly under-represented in large-scale genomic mapping projects such as TCGA datasets. We retrieved 10 Stage IIIA CRC cases, matched these to 16 Stage 1 CRC cases (T1 depth without lymph node metastasis) and carried out deep sequencing of 409 genes using the IonTorrent system. Tumour mutational burdens (TMB) ranged from 2.4-77.2/Mb sequenced. Using mean TMB as a cut-point to define groups, TMB-low (TMB-L) specimens showed higher frequency of *KRAS* and *TP53* mutations in Stage IIIA compared to Stage I, consistent with TCGA data. TMB-High (TMB-H) specimens consisted of both microsatellite instable high (MSI-H) and microsatellite stable (MSS) genotypes. Comparison of TMB-H with TMB-L groups revealed clear differences in mutations of *ATM, KDM5C, PIK3CA* and *APC. APC* was less frequently mutated in the TMB-H group although variant composition was more diverse. Variants in *ATM* were restricted to the TMB-H group, and in four of five MSS specimens we observed the co-occurrence of mutations in homologous recombination repair (HRR) genes in either two of *ATM, CDK12, PTEN* or *ATR*, with at least one of these being a truncating mutation. No MSI-H specimens carried nonsense mutations in these genes. These findings add to our knowledge of early T stage CRC and highlight a potential therapeutic vulnerability in the HRR pathway of TMB-H MSS CRC.

## Introduction

Colorectal cancer (CRC) maintains a high worldwide disease burden as the third most common malignancy and the second leading cause of cancer mortality [1]. In 2020, CRC accounts for 10% of global cancer incidence and 9.4% of cancer deaths [2]. Molecular studies have revealed two major and distinct genetic/epigenetic pathways for sporadic CRC development [3]. The conventional adenoma-carcinoma pathway is driven by APC/beta-catenin/Wnt disruption, often coupled with *KRAS* oncogene activation resulting in chromosomal instability. CRC arising in this pathway is common and these tumours have proficient DNA mismatch repair mechanisms and are characterised as microsatellite stable (MSS) [4]. In contrast, CRC arising from the serrated pathway is characterised by high levels of CpG island methylator phenotype (CIMP) due to epigenetic silencing of the *MLH1* DNA mismatch repair gene [5]. Deficient mismatch repair and high microsatellite instability (MSI-H) leads to the accumulation of many hundreds of mutations fuelling carcinogenesis. Large-scale genomic profiling projects such as The Cancer Genome Atlas (TCGA) have contributed significantly more details as to the identity and frequencies of driver and other mutations in CRC [6]. For example, review of TCGA-COAD dataset revealed *APC* (77%), *TP53* (58%), *KRAS* (44%), *MUC16* (38%), *PIK3CA* (31%), and *OBSCN* (30%) as the top six mutated genes in sporadic colon adenocarcinoma (https://portal.gdc.cancer.gov/projects/TCGA-COAD).

However, not all AJCC/TNM stages of CRC [7] are equally represented in TCGA data. One of the most under-represented is Stage IIIA, diagnosed in approximately 10% of all locally advanced CRC (LAC), and signifies the earliest stage of LAC. The combined TCGA-COAD and TCGA-READ datasets consist of only 12 patients with Stage IIIA CRC, greatly limiting our knowledge on the molecular drivers underlying tumour development. Stage IIIA CRC (T1-T2 N1/N1c M0 or T1 N2a M0) is defined by lymph node metastasis (LNM), despite the primary lesion having limited invasive depth no greater than into the muscularis propria (T1-T2). Stage IIIA patients have good prognosis, although a considerably higher metastatic relapse rate compared with Stage I tumours [8,9] which show similar invasive depth, but no lymph node metastasis (T1-2 N0 M0). Determining the likelihood of LNM in T1-T2 cancers remains an important unmet clinical need, especially from endoscopic polypectomy specimens. This is because many patients with a T1-T2 diagnosis post-polypectomy, undergo additional radical surgery where the subsequent diagnosis of LNM is reported to be as low as 8-16%, and these patients are exposed to potential post-operative complications [10,11]. Using proteomics we have previously shown that stage IIIA CRC are enriched in protein regulators of epithelial-mesenchymal transition (EMT), and proposed that this may be useful in identifying high risk polypectomy specimens [12]. To complement this study, molecular genomic analysis focusing on stage IIIA CRC is needed to establish mutational profiles, risk of LNM, and further improve prognostication and treatment approaches.

Stage IIIA CRC is infrequent. We undertook massively parallel sequencing on our stage IIIA CRCs to establish their common mutational profiles and compared this to a matched Stage I CRC cohort. By focusing on early T stage CRCs, we made interesting observations on mutational burden and pathway defects in MSS and MSI-H CRC that has not been widely reported.

## Materials and Methods

### Specimens

The study was approved by the Northern Sydney Local Health District Human Research Ethics Committee (RESP/18/248). FFPE colorectal adenocarcinoma specimens from patients who underwent surgical resection between 2007-2018 were acquired from the Department of Anatomical Pathology, Royal North Shore Hospital (Sydney, Australia). Lynch syndrome cases were excluded from the study. We utilised specimens where sufficiently high-quality DNA could be obtained from formalin fixed paraffin embedded (FFPE) blocks resulting in 16 stage I and 10 stage IIIA cases based on their synoptic pathological staging according to 8^th^ edition AJCC/TNM staging system [7]. Mismatch repair protein (MMRp) immunoexpression and MSI status were examined as part of routine clinical histopathology reporting, showing four cases were MSI-H cancers. Combined BRAFV600E/MMRp status was determined and showed two MSS/BRAF mutant cases (2/26), four MSI-H/BRAF mutant cases (4/26), and twenty MSS/BRAF wild-type cases (20/26).

### Next Generation Sequencing

Hematoxylin and eosin stained tissue sections from FFPE tumour samples first were reviewed by a qualified pathologist (M.A.) and tumour region was marked out on the slides. Extraction of tumour area was achieved by macrodissection. DNA/RNA recovery followed the manufacturer’s protocol of AllPrep DNA/RNA FFPE Kit (QIAGEN).

#### Ion AmpliSeq™ Comprehensive Cancer Panel

Targeted sequencing was achieved by Ion AmpliSeq™ Comprehensive Cancer Panel (ThermoFisher Scientific), which consisted of 409 key cancer-related genes. Library preparation for each specimen was performed using the Ion Ampliseq Library Kit 2.0 (ThermoFisher Scientific) according to the manufacturer’s instructions. The prepared libraries were partially digested and phosphorylated using the FuPa reagent, ligated to different barcode adapters using the Ion Xpress Barcode Adapters Kit (ThermoFisher Scientific), then purified. The purified libraries were quantified using the Ion Library TaqMan Quantitation Kit (ThermoFisher Scientific).

Data analyses for variant calling (SNVs/multi-nucleotide variants [MNVs], and indels) and annotation were performed in Ion Reporter software version 5.14 (**https://ionreporter.thermofisher.com/ir/**) following workflows of Oncomine Tumour Mutation Load -w3.1 – DNA and Oncomine Variant Annotator v3.0 plug-in using default settings. Filter chain of Oncomine Extended (version 5.12) was launched to identify somatic variants. Human genome build 19 was used as reference in alignment. A minimum sequencing depth of 500X was considered as adequate sequencing depth, and variant allelic fraction (VAF) of 5% was used as a cut-off for positive variants. Gene/variant summaries and plots were prepared by maftools [13].

### Retrieval of TCGA datasets

TCGA COAD and READ projects were used as an independent validation data source. COAD and READ data were downloaded from GDC data portal (https://portal.gdc.cancer.gov/) and inclusive criterion were disease type of “adenomas and adenocarcinomas” and AJCC pathologic stages of “stage I” and “stage IIIA”. In total 96 stage I and 12 stage IIIA were available.

## Results

### Robust Deep Targeted sequencing of early T stage colorectal cancer

TCGA COAD and READ datasets report molecular genomics on 550 cases, with only 12 cases defined as Stage IIIA CRC. Over the past decade in our institution, Stage IIIA CRC was diagnosed in approximately 4% of all surgical cases. We identified 10 Stage IIIA cases where FFPE blocks were available and DNA was recovered with sufficient quality for deep sequencing. A matched cohort of 16 Stage I CRC was assembled for comparison as early T stage tumours without lymph node metastasis. Clinicopathological characteristics of the cohort used in this study is shown in Table 1. Stage IIIA patients were younger, and by definition, had nodal involvement or sub-serosal deposits as the only significant variables identified in the cohort.

**Table 1.**
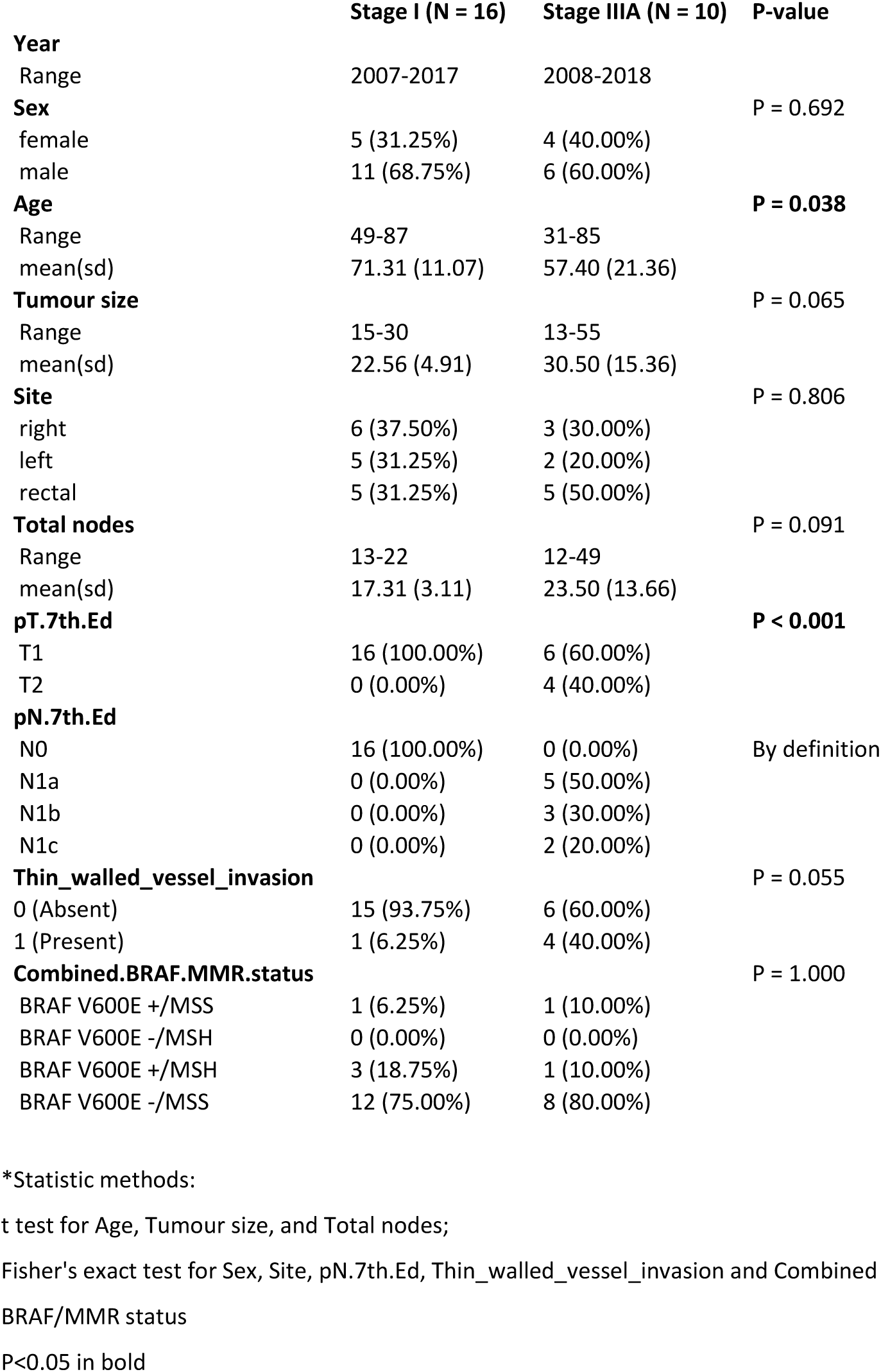
Clinical characteristics of Stage I and Stage IIIA CRC samples.

DNA from 26 FFPE CRC specimens were analysed using the IonTorrent AmpliSeq™ Comprehensive Cancer Panel, which targeted the exons of 409 tumour suppressor genes and oncogenes. One sample was excluded from data analysis as sequencing was below the quality threshold. For the remaining 25 samples, the range of mean sequencing depth was 876 – 2253-fold; mapped reads were 15,073,849 – 35,403,831; uniformity was between 86.59% - 96.25%; on-target reads were 93.66% - 99.19%. dbSNP concordance rate was 0.836-0.992. These metrics confirm robust deep sequencing data quality.

Tumour mutation burden (TMB) varied from 2.4 per Mb to 77.2 per Mb, mean 16.5 per Mb. Somatic mutation characteristics and relative clinical features are shown in Figure 1. We classified seven samples with TMB > 16.5 per Mb into a TMB-High (TMB-H) group. In these seven samples with high mutational burdens (Figure 1), two (28.6%) had high microsatellite instability (MSI-H), while the five remaining TMB-H specimens were considered to be microsatellite stable (MSS). Two additional MSI-H samples had TMB of 14.4 and 8.8. To enable further analysis we combined all IHC confirmed MSI-H and TMB-H cancers into a single group of nine hypermutated specimens. (Mean TMB-H group 35.2/Mb, Mean TMB-Low group 5.9/Mb)

**Figure 1.**
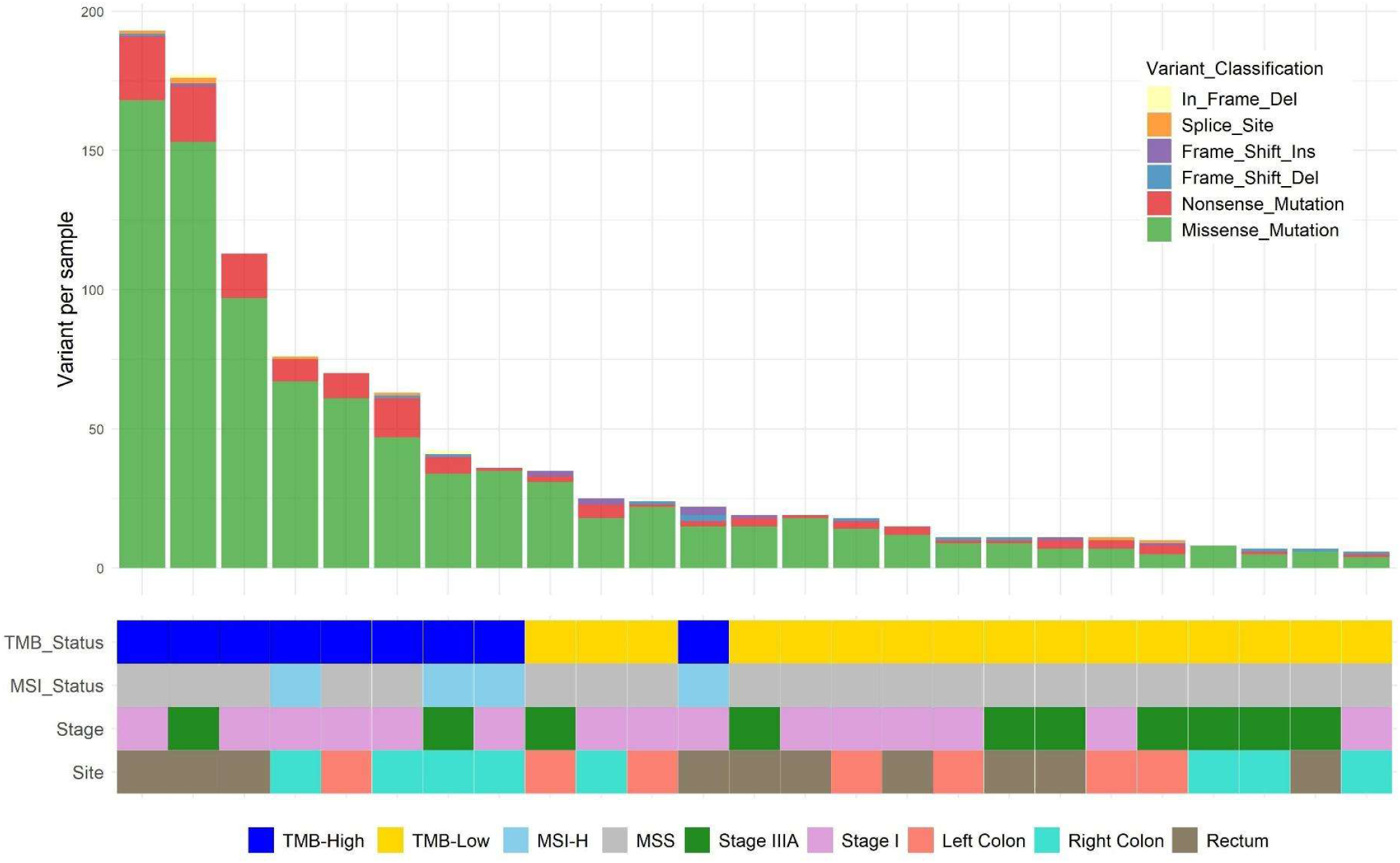
Summaries of variant profiling in TMB-High and TMB-Low groups. Mean TMB 16.5 per Mb.

### No stage-related mutational differences between Stage IIIA vs Stage I in TMB-Low group

To further elucidate the molecular differences between Stage IIIA and Stage I CRC, we focused on the TMB-L group consisting of eight Stage I and eight Stage IIIA samples. In the TMB-H group there were insufficient Stage IIIA cases for a meaningful analysis. In comparing all 409 cancer-related genes, *KRAS* (62.5% v 12.5% in stage IIIA v stage I) and *TP53* (85.7% v 50%) showed a higher mutation rate in stage IIIA compared to Stage I cancers, although this did not reach statistical significance (Table 2). No stage-related difference was observed in mutational frequencies of *APC, PIK3CA, FBXW7*, and *SMAD4* CRC driver genes. We compared our findings with TCGA COAD and READ cohorts, adopting the TCGA defined threshold for discriminating hypermutators from non-hypermutators (i.e. >12 per 10^6^ (total mutations > 728) [6]). As shown in Supplementary Table S1 comparison of 69 Stage I and 12 Stage IIIA TCGA specimens [6] showed no significant difference in gene frequencies, consistent with our observations.

**Table 2.** Statistical analysis* on differential distributed genes between stage IIIA and stage I.

| Gene<br>symbol | Stage IIIA<br>(freq%) | Stage I<br>(freq%) | pval | Odds<br>ratio | Confidence<br>Interval .Upper | Confidence<br>Interval.lower | adjPval |
| --- | --- | --- | --- | --- | --- | --- | --- |
| KRAS | 5 (62.5%) | 1 (12.5%) | 0.12 | 9.76 | 625.22 | 0.68 | 0.24 |
| TP53 | 7 (87.5%) | 4 (50%) | 0.28 | 6.16 | 391.63 | 0.42 | 0.38 |
| APC | 7 (87.5%) | 7 (87.5%) | 1 | 1 | 89.51 | 0.01 | 1 |

\*Performed fisher exact test on gene frequencies of two groups; P value was adjusted by fdr.

We next used Oncoplots to compare frequent gene variants in TMB-L cohort with findings from the TCGA COAD and READ non-hypermutated cohort (Figure 2). Taking our top 10 genes we noted consistency in the top three genes *APC* (88% v 85%), *TP53* (69% v 57%) and *KRAS* (38% v 43%), but observed higher frequencies of the remaining seven genes (*TAF1* (31% v 1%), *FBXW7* (31% v 11%), *NTRK1* (25% v 0%), *ARID2* (25% v 2%), *RET* (25% v 2%), *TAF1L* (25% v 4%), and *USP9X* (19% v 4%)) compared with the TCGA cohorts (Figure 2). The reciprocal analysis using the top 10 genes from TCGA cohorts is presented in Supplementary Figure S1. *APC* (85%), *TP53* (57%), *KRAS* (43%), *PIK3CA* (20%), *SYNE1* (19%), *LRP1B* (12%), *FBXW7* (11%), *CARD11* (9%), *ATM* (9%) and *NRAS* (9%) are the most frequent 10 genes in TCGA COAD and READ. We detected fewer variants in *PIK3CA* (6% v 20%) and *ATM* (0% v 9%) compared with TCGA cohorts, while the frequencies of other genes were consistent. It is noteworthy that only 12 specimens of Stage IIIA exist in the TCGA cohort and the small cohort sizes of our data and the TCGA cohort is a likely contributing factor to the disparate frequencies observed for some genes. Furthermore, our detection of more variants is likely attributed to the robustness of sensitive deep sequencing method we used, compared with WXS used in TCGA projects.

**Figure 2.**
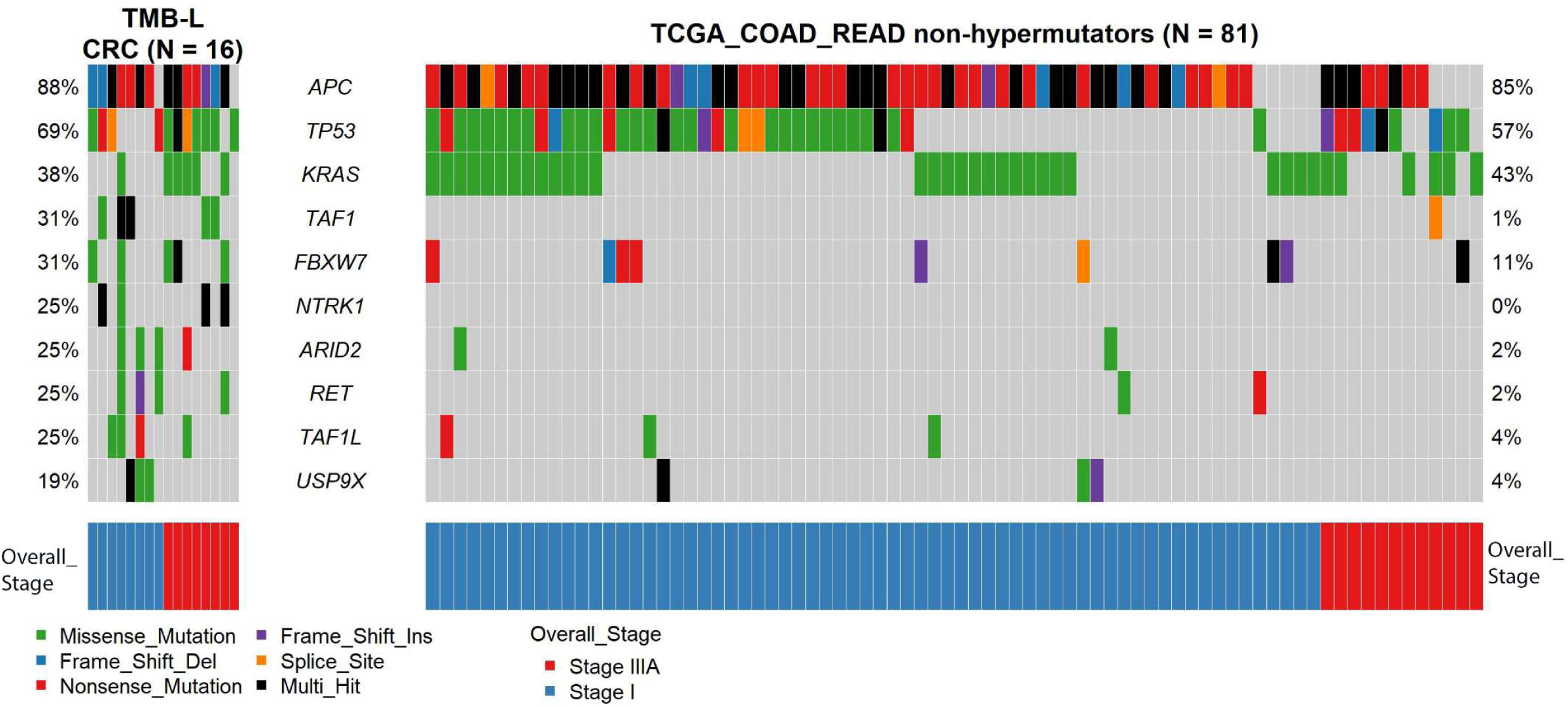
Parallel view of top ten mutated genes in CRC TMB-Low samples and non-hypermutators TCGA COAD-READ cohort specimens.

### Mutational spectrum of TMB-H and TMB-L CRC

This study allowed us to compare TMB-H and TMB-L groups of early T stage CRC where we observed different ranking of the top mutated genes. *ATM* (67%), *KDM5C* (56%), *APC* (44%) and *PIK3CA* (56%) were common cancer driver genes found in TMB-H samples, while in TMB-L groups *APC* (88%), *TP53* (69%), and *KRAS* (38%) were the most frequent driver genes (Table 3). We checked the frequencies of the above genes in TCGA COAD-READ cohort and found similar distribution trends of dominant driver genes in TCGA non-hypermutators and hypermutators, though some discrepancies in gene frequencies were evident (Table 3).

**Table 3.** Different distribution of cancer driver genes in Stage I and IIIA hypermutated and non-hypermutated CRC.

| RefSeq<br>Gene | TMB-H (n=9) | TMB-L (n=16) | TCGA COAD/READ<br>hypermutated<br>(n=10) | TCGA COAD/READ non-hypermutated (n=81) |
| --- | --- | --- | --- | --- |
| ATM | 67% | 0 | 60% | 9% |
| KDM5C | 56% | 6% | 10% | 1% |
| PIK3CA | 56% | 6% | 40% | 20% |
| APC | 44% | 88% | 40% | 85% |
| TP53 | 44% | 69% | 20% | 57% |
| KRAS | 44% | 38% | 10% | 43% |

In our study, although *APC* was frequently mutated in both TMB-H and TMB-L, it was less frequently mutated (44% vs. 88%) in the TMB-H group. Nonetheless, when *APC* was mutated in TMB-H cancers, a more diverse composition of variant types was observed: three missense, three nonsense, one frameshift indels, and one splice-site variant. Conversely, among 16 TMB-L samples, none of the *APC* alterations was missense/synonymous. Instead, we observed potentially high impact deleterious *APC* mutations consisting of 14 nonsense mutations and five frameshift indels. Lollipop plot (Figure 3) reveals the distributions of *APC* variants in the context of functional protein domains. TMB-H CRC showed a more even distribution of *APC* variants dispersed over the codons of 516 – 2620; while TMB-L CRC contained 58% (11/19) of amino acid changes located in the mutation cluster region (MCR, codons 1282–1581), which is in the central part of the *APC* coding frame involved in β-catenin downregulation [14]. Clustering of *APC* variants (49%) also can be observed in MCR of TCGA COAD-READ non-hypermutators (Supplementary Figure S2).

**Figure 3.**
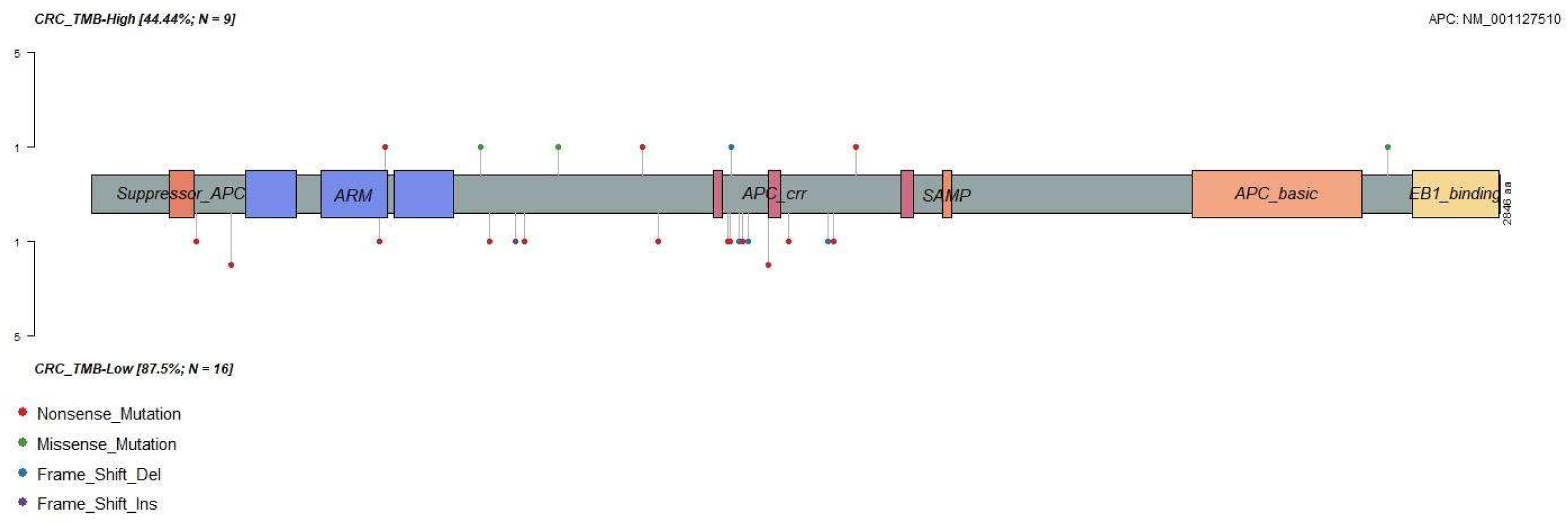
Lollipop plot shows different distributions of *APC* variants between TMB-High and TMB-Low CRC. *APC* variants present in 44% of nine TMB-H CRC cases with an even distribution from codon 516 – 2620. Conversely in TMB-Low group of 16 CRC cases, *APC* has a higher mutational rate 87.5% and 58% of variants is enriched in the mutation cluster region (MCR, codons 1282– 1581) in the central part of *APC*.

### Deficiency of homologous recombination repair in TMB-H CRC

Interestingly, variants in *ATM* were only present in the TMB-H tumours. Significant difference of *ATM* distribution was also observed in the analysis of early T stage TCGA hypermutated (60%) and non-hypermutated groups (9%) (p = 0.0004) (Table 3). According to the TCGA publication that analysed all stages of CRC [6], *ATM* did not appear as a top ranked recurrently mutated gene in either hypermutated (rank 38) or non-hypermutated (rank 20) cohorts. Our dataset highlighted *ATM* as a potential indicator for TMB-H or hypermutated CRC, evident in early T stage cancers here, and consistent with the data available from TCGA early T stage CRC. That drove our attention to the homologous recombination repair (HRR) pathway, one of the most important cellular pathways for repair of double strand DNA breaks, which has been associated with cancer predisposition and increased sensitivity to chemotherapeutic agents that cause double strand breaks [15]. On the AmpliSeq panel, 13 HRR pathway genes are present: *ATM, PTEN, BRIP1, ATR, PALB2, MRE11A, RAD50, CHEK1, CHEK2, FANCD2, FANCA, XRCC2* and *CDK12*. The Figure 4 Oncoplot shows the co-occurrence of multiple mutations in either two of *ATM, CDK12, PTEN* and *ATR* in TMB-H MSS CRC identified here for the first time (Supplementary Table S2). In our data, four out of five TMB-H MSS CRC carry at least two truncating mutations or multi-hit deleterious lesions in these four HRR genes. The remaining TMB-H MSS case displays a missense variant in *FANCD2* only, which is predicted to be pathogenic, suggesting a distinct mechanism of high mutational burden rise. Importantly, none of the MSI-H CRC showed HRR gene truncating mutations.

**Figure 4.**
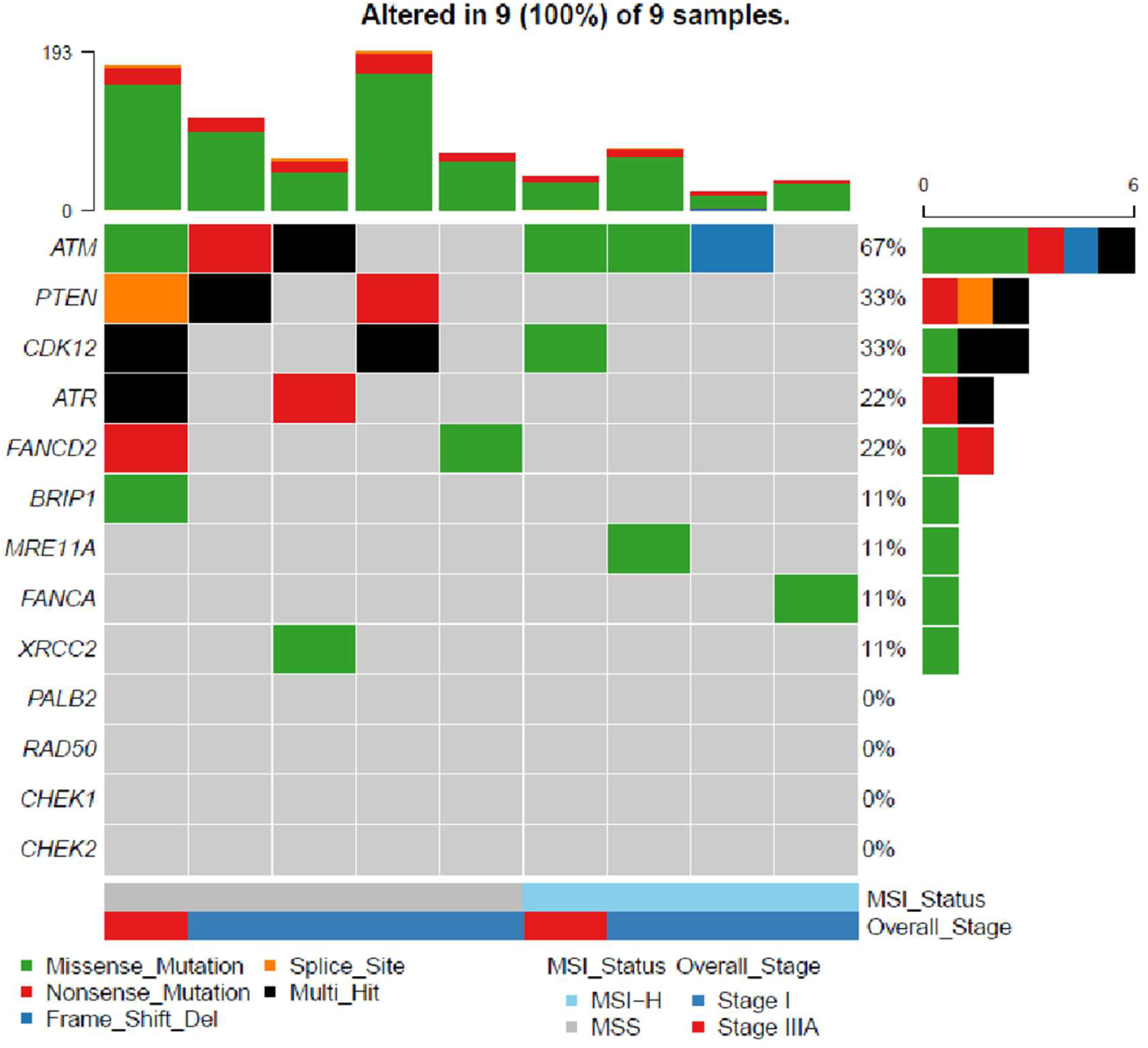
HRR gene co-occurrence in TMB-H MSS CRC specimens

### Oncogenic Pathway analysis

Since our study used targeted sequencing of key cancer genes, we evaluated frequent alterations in ten canonical signaling pathways as described in previous TCGA reports [16] (Figure 5). As expected, TMB-H samples harboured more altered oncogenic pathways due to higher mutation load. RTK-RAS signalling pathway was the most frequently affected pathway among early T stage cancers. TCGA project also identified the RTK-RAS signaling pathway as the most frequently altered oncogenic network in CRC MSI-POLE group, second most frequent in CRC GS (genomically stable), and third most frequent in CRC CIN (chromosomal instability) [9]. Other high frequency affected pathways in the TMB-H cancers were PI3K, NOTCH, TP53 and WNT, whereas less frequently affected pathways were observed for PI3K and NOTCH in TMB-L cancers. Discrepancies were observed for HIPPO and MYC pathways in TMB-L cancers due to a lack of genes from those two pathways present on AmpliSeq panel.

**Figure 5.**
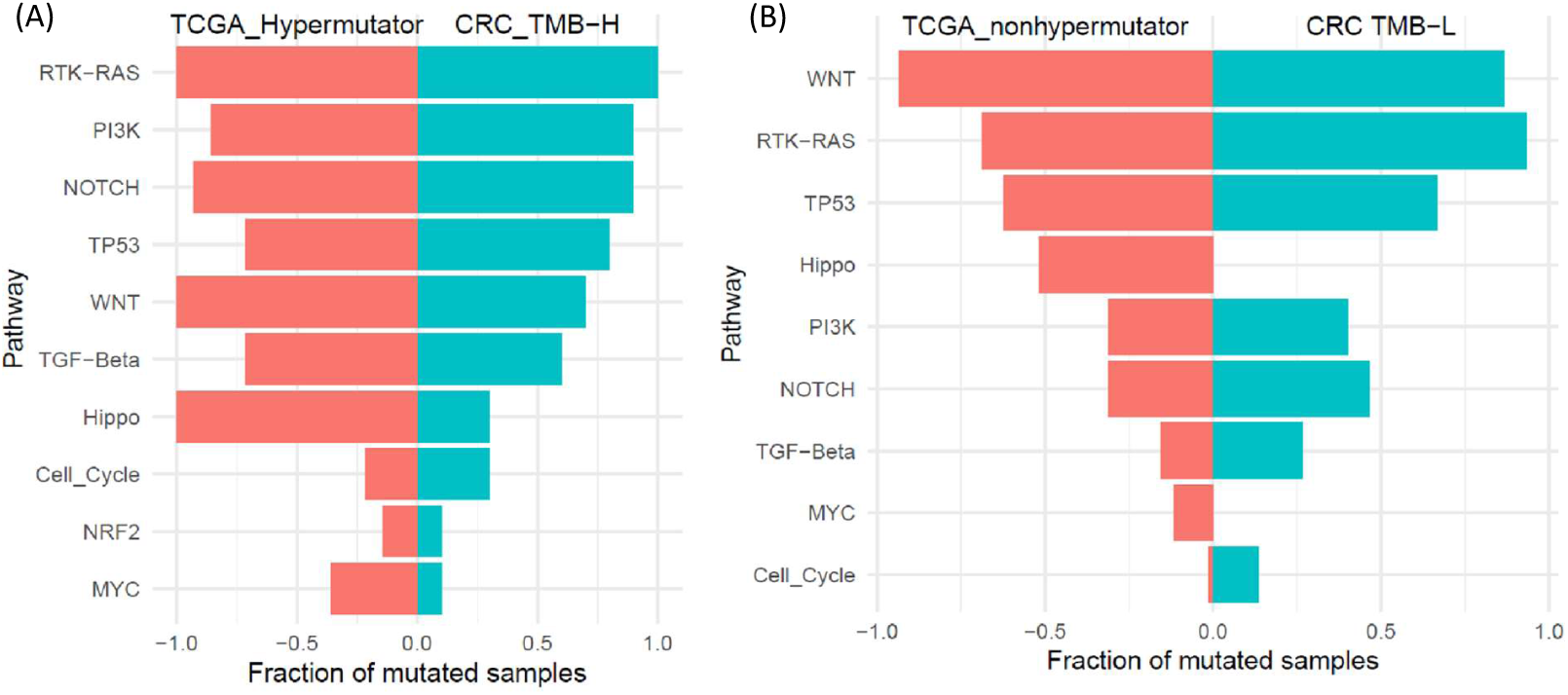
Pathway enrichment analysis in (A) TMB-H group compared with TCGA-COAD-READ hypermutator group, (B)TMB-L group compared with TCGA COAD-READ non-hypermutated cohort.

## Discussion

In this study we set out to profile common cancer-related mutations that are seen in early T stage CRCs, with a particular interest in Stage IIIA CRC given the limited genomic data available for this disease group. We adopted in-depth massively parallel sequencing using the Ion AmpliSeq™ Comprehensive Cancer Panel which contained 409 key cancer-related genes and contributed an additional 80% of genomic profiles for Stage IIIA CRC cases beyond that reported by TCGA. While recognising the small study size, our analysis did not reveal any differentially distributed gene mutation that would stratify LNM positive T1-2 patients from Stage I CRC patients.

The use of massively parallel sequencing enabled calculation of TMB, enabling us to define TMB-H and TMB-L for separate analyses. A key observation arising was the enrichment of some MSS CRC in the TMB-H group. Hypermutated and MSI-H CRC are predictive factors for favourable response to immunotherapy and improved survival [17]. In our study, we observed seven tumours with high mutational burden (2 MSI-H and 5 MSS) and two additional MSI-H tumours with TMB below the mean (Figure 1). Interestingly, seven out of nine hypermutated samples (78%) originated from Stage I patients, whilst a more balanced composition of eight stage I vs eight stage IIIA was found in the non-hypermutated group. Four hypermutators were in the right side of the colon, one from the left colon and four were rectal cancers. Hu et al. examined hypermutated CRC samples in TCGA data and discovered similar distribution as our data with 77% of TCGA hypermutated CRC samples derived from stage I/II patients and 80% were located in the right side of the colon [18].

Our study points out the promising role of homologous recombination repair (HRR) pathway gene mutations, specifically the co-occurrence of *ATM* with *CDK12, PTEN* or *ATR* in identifying hypermutated MSS CRC; a finding we confirmed is observable in the larger TCGA-COAD/READ Stage I/IIIA dataset (Supplementary Figure S2). This has potential translational significance as HRR deficiency (HRD) is targetable with PARP inhibitors to exploit synthetic lethality, as is the case with BRCA1/2 ovarian cancer [19]. Exploiting such a vulnerability in MSS CRC that carry “BRACness” genetics [20] would be a paradigm shift, where chemotherapy has remained the only treatment option for advanced MSS CRC for decades.

A significant barrier to targeting HRD in all cancers has been establishing best practice to identify the characteristics of HRD tumour status. In clinical practice, it is currently infeasible to use WES or deep panel sequencing as we applied here to determine TMB and find hypermutated MSS. Identifying BRACness CRC may be feasible using a targeted panel of HRR genes as applied in other cancer settings [21].

Lee et al. analysed hypermutated MSS CRC with *BRCA1/2* somatic truncations using all stages CRC from TCGA PanCancer study as discovery set (N = 21) and metastatic CRC subset of MSK-IMPACT (Memorial Sloan Kettering Cancer Center [MSKCC]) as validation cohort (N = 41). They concluded that somatic truncations of *BRCA1/2* in combination with other HRR genes can identify MSS hypermutators with or without known pathogenic exonuclease domain mutations in *POLE* [22].

Interestingly, in analysis of true positive rates to identify MSS hypermutant CRC, Lee et al predicted the best performed three individual genes were *PTEN, ATM* and *ATR*. Since *BRCA1/2* are not included on the Ion AmpliSeq™ Comprehensive Cancer Panel, we could not confirm this in our dataset of early T stage CRC, so we examined their frequencies in TCGA data (Supplementary Figure S3) and found the mutational frequencies of BRCA2 and BRCA1 were only 30% and 20% respectively in hypermutant early-T stage CRC. In our study we report the co-occurrence of truncating mutations in *ATM, CDK12, PTEN* or *ATR* is present in 80% of MSS CRC with TMB-H and absent from TMB-L MSS cancers. When applied to the TCGA data the co-occurrence of *ATM* with another HRR gene outperformed use of *BRAC1/2*, yielding 60% detection rate. Discrepancy on the weight of *BRCA1/2* in predicting hypermutant MSS cases might be related to advanced disease stages dominating the report of Lee et al [22], while our study focused on early T stage CRC.

While sample size of our study and that reported in the TCGA for early T stage CRC with MSS is small our findings provide a testable pathway for the potential exploitation of HRD in hypermutant MSS CRC. We believe this is the first report that shows deleterious *ATM* mutations significantly enriched in hypermutated early T stage CRC. Future large prospective study will be required to demonstrate the translational feasibility of our finding as promising biomarkers to identify suitable patients for treatments targeting HRD.

## Conclusions

This study has significantly increased the molecular genomics knowledge base for Stage IIIA CRC. In MSS Stage IIIA CRC which carried low TMB we observed higher frequency of *KRAS* and *TP53* mutations compared to Stage I CRC. We made the interesting observation in MSS CRC with high TMB that these tumours are defined by non-synonymous variants affecting genes of the HRR pathway, commonly *ATM*, but also others. Moreover, these MSS tumours harboured co-occurring mutations in HRR genes including at least one truncating mutation. This finding may be significant if observed in advanced CRC as it highlights a potential vulnerability which may be exploited therapeutically using the combination of agents that cause DNA single strand breaks and PARP inhibitors.

## Data Availability

All data produced in the present study are available upon reasonable request to the authors

## Supplementary Material

**Table S1**. Gene frequency differences amongst 69 Stage I and 12 Stage IIIA CRC specimens from TCGA COAD-READ projects.

**Table S2**. Gene variants identified in TMB-H MSS CRC samples.

**Figure S1**. Oncoplot showing the most frequently mutated genes from non-hypermutators in TCGA COAD-READ project compared with TMB-low CRC in this study.

**Figure S2**. Lollipop plot shows distributions of *APC* variants in hypermutated and non-hypermutated TCGA COAD-READ cohorts.

**Figure S3**. Oncoplot of HRR genes in the ten hypermutators of TCGA COAD-READ cohort.

## Acknowledgement

The authors acknowledge the technical assistance of (Dr. Cali Willet and Dr. Tracy Chew) of the Sydney Informatics Hub, a Core Research Facility of the University of Sydney.

## Author contributions

Conceptualization, J.L., P.S., A.F.E., and M.P.M.; Software, J.L.; Investigation, J.L., P.S., B.C.Y.T., M.S.A.; Resources, A.J.G.; Data Curation, J.L and M.P.M.; Writing – Original Draft Preparation, J.L.; Writing – Review & Editing, M.S.A., B.C.Y.T., A.F.E., M.P.M.; Supervision, A.J.G., M.P.M.; Funding Acquisition, J.L., A.F.E., and M.P.M. All authors have read and agreed to the published version of the manuscript.

## Institutional Review Board Statement

The study was conducted according to the guidelines of the Declaration of Helsinki, and approved by the Human Research Ethics Committee of Northern Sydney Local Health District (RESP/18/248).

## Informed Consent Statement

The approved protocol provided for a waiver of patient consent as specimens were collected for the purpose of disease treatment and the impracticality and distress of contacting patients many years following such procedures.

## Data Availability Statement

All data produced in the present study are available upon reasonable request to the authors

