## Supplementary material for "Deep sequencing of early T stage colorectal cancers reveals disruption of homologous recombination repair in microsatellite stable tumours with high mutational burdens": Figure S1

**Supplementary Figure S1**. Oncoplot showing the most frequently mutated genes* from non-hypermutators in TCGA COAD-READ project compared with TMB-low CRC in this study.

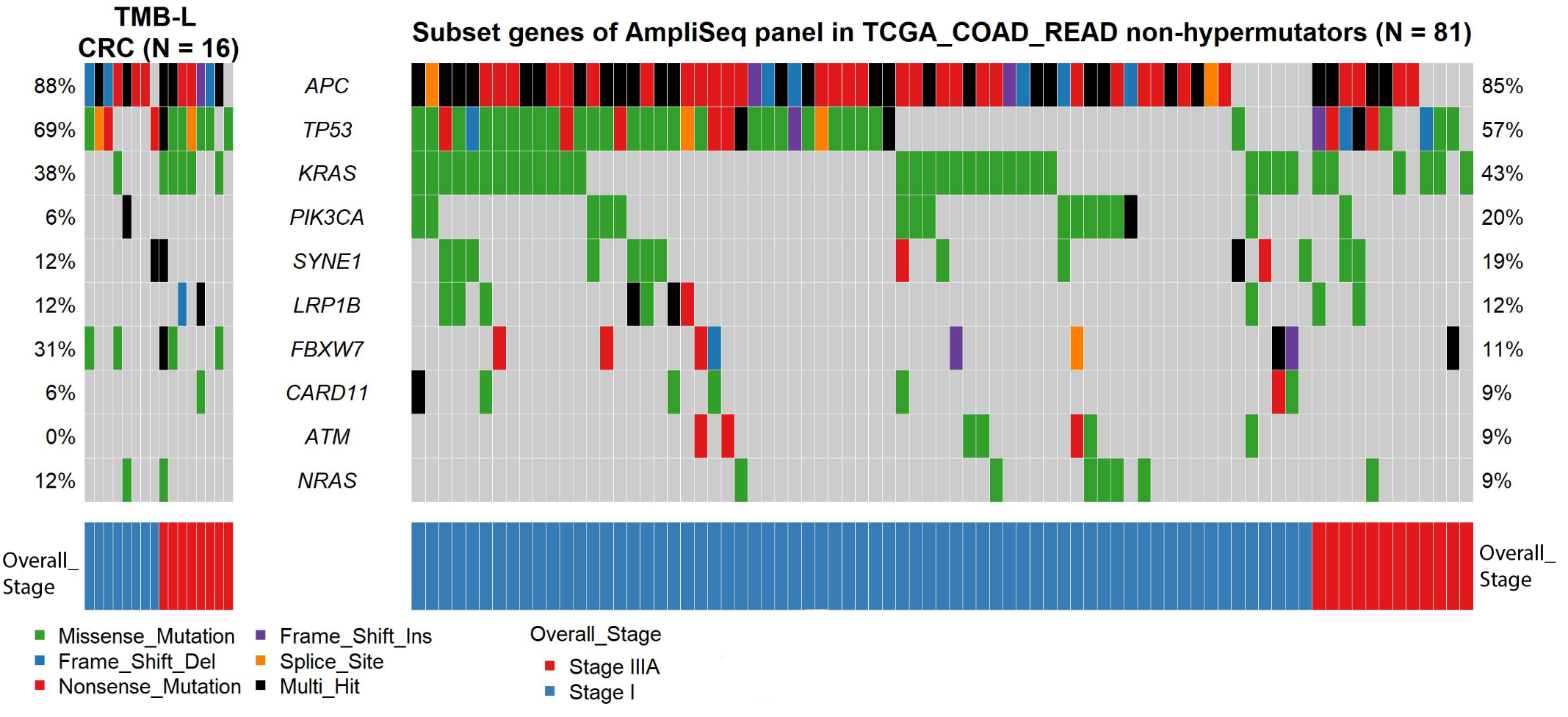

*Considering mutation files of TCGA-COAD-READ projects were generated via WXS, we hereby focused on the subset of 409 genes on the Ion AmpliSeq™ Comprehensive Cancer Panel in order to do meaningful comparison between our data and TCGA-COAD-READ data. These figures demonstrate observations between our study and TCGA are mostly consistent.
