## Supplementary material for "Deep sequencing of early T stage colorectal cancers reveals disruption of homologous recombination repair in microsatellite stable tumours with high mutational burdens": Figure S2

**Supplementary Figure S2**. Lollipop plot shows distributions of APC variants in hypermutated and non-hypermutated TCGA COAD READ cohorts

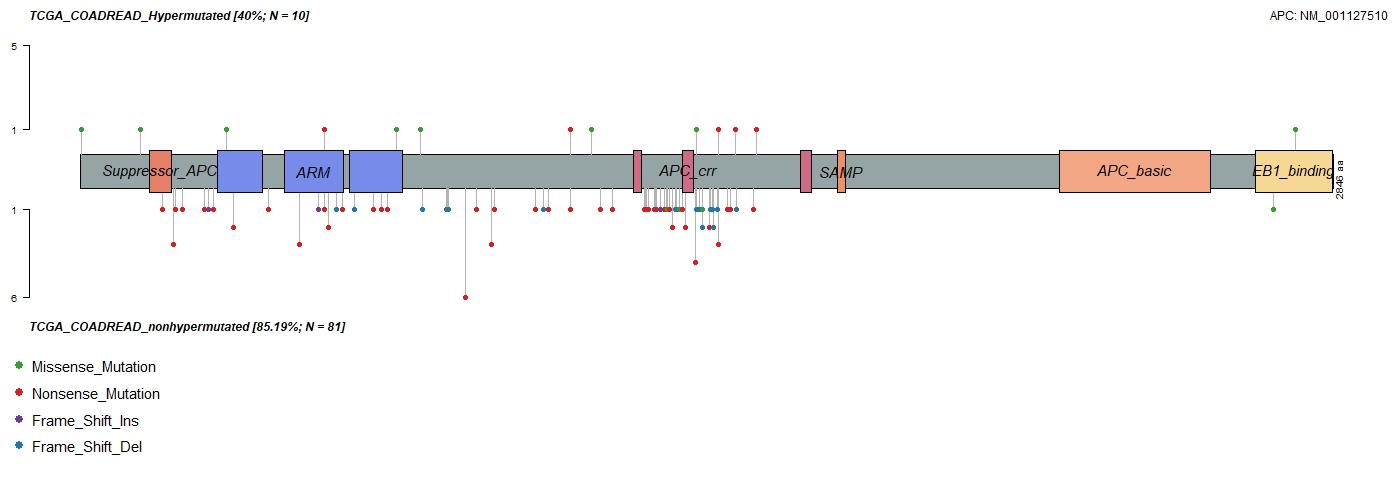
