## Supplementary figures and images for "Deep sequencing of early T stage colorectal cancers reveals disruption of homologous recombination repair in microsatellite stable tumours with high mutational burdens"

### Figure S3

**Supplementary Figure S3**. Oncoplot of HRR genes in the ten hypermutators of TCGA COAD-READ cohort

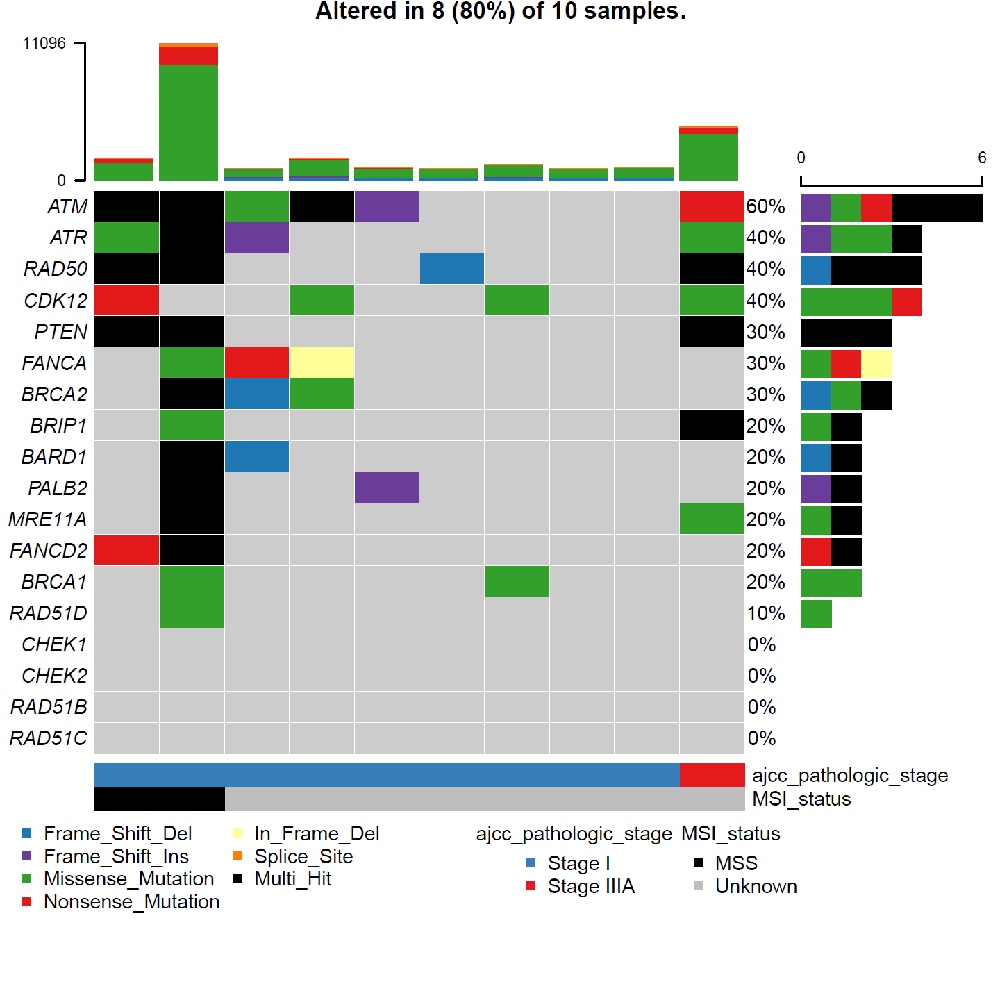
